# Burden of Latent Tuberculosis Infection among Healthcare Workers of Primary Health Centres and its Association with Clinical Laboratory Parameters

**DOI:** 10.1101/2024.09.27.24314525

**Authors:** Sivaprakasam T. Selvavinayagam, Ganga Sankar, Yean K. Yong, Sathish Sankar, Ying Zhang, Hong Y. Tan, Pachamuthu Balakrishnan, Amudhan Murugesan, Manivannan Rajeshkumar, Asha Frederick, Masilamani Senthil Kumar, Paulkanraj PriyaRaj, Jesudoss Prabhakaran, Pattusamy Sangeetha, Pasupathy Arunpathy, Rajamani Charu, Nagarajan Muruganandam, Deepak M. Sakate, Deepak Jayakumar, Prabu Dhandapani, Parthiban Rudrapathy, Vijayakumar Velu, Marc Emmenegger, Marie Larsson, Esaki M. Shankar, Sivadoss Raju

**Affiliations:** Directorate of Public Health and Preventive Medicine, DMS Campus, Teynampet, Chennai, Tamil Nadu, India; State Public Health Laboratory, Directorate of Public Health and Preventive Medicine, DMS Campus, Teynampet, Chennai 600 006, Tamil Nadu, India; Laboratory Center, Xiamen University Malaysia, 43900 Sepang, Selangor, Malaysia; Kelip kelip! Center of Excellence for Light Enabling Technologies, Xiamen University Malaysia, Sepang, Selangor, Malaysia; Department of Microbiology, Centre for Infectious Diseases, Saveetha Dental College and Hospitals, Saveetha Institute of Medical and Technical Sciences, Saveetha University, Chennai 600 077, Tamil Nadu, India; Chemical Engineering, Xiamen University Malaysia, Sepang, Malaysia; School of Traditional Chinese Medicine, Xiamen University Malaysia, Sepang, Malaysia; Department of Research, Meenakshi Academy of Higher Education and Research (MAHER), Chennai 600 078, India; Department of Microbiology, Government Theni Medical College and Hospital, Theni 625 512, India; Indian Council Medical Research - Regional Medical Research Centre, Port Blair 744 103, Andaman and Nicobar Islands, India; Department of Statistics and Applied Mathematics, Central University of Tamil Nadu, Thiruvarur, Tamil Nadu, India; Department of Microbiology, Dr. ALM Post Graduate Institute of Basic Medical Sciences, University of Madras, Chennai, Tamil Nadu, India; Microbiology Division, Department of Clinical Laboratory Services and Translational Research, Malabar Cancer Centre (Post Graduate Institute of Oncology Sciences and Research), Thalassery 670 103, Kerala, India; Department of Pathology and Laboratory Medicine, Emory University School of Medicine, Division of Microbiology and Immunology, Emory National Primate Research Center, Emory Vaccine Center, Atlanta, GA 30329, USA; Institute of Neuropathology, University Hospital Zurich, Zürich, Switzerland; Division of Medical Immunology, Department of Laboratory Medicine, University Hospital Basel, 4031 Basel, Switzerland; Division of Molecular Medicine and Virology, Department of Biomedical and Clinical Sciences, Linköping University, 58 185 Linköping, Sweden; Infection and Inflammation, Department of Biotechnology, Central University of Tamil Nadu, Thiruvarur 610 005, India

**Keywords:** Latent tuberculosis, Healthcare worker, Primary health centre, TB, India, IGRA, Interferon Gamma Release Assay

## Abstract

**Objectives:** Healthcare workers (HCWs) are at high risk of latent tuberculosis infection (LTBI) due to their continued occupational exposure. The incidence of LTBI among the HCWs has seldom been investigated. Primary health centres (PHCs) provide effective and affordable medical care largely for the local population. The HCWs of PHCs who are likely to have increased occupational exposure have an increased risk of reactive as well as LTBI, and contribute to overall TB transmission than the general population.

**Methods:** A cross-sectional study (March–April 2024) was carried out to assess the burden of LTBI among the HCWs of PHCs (n=64) across Chennai, India. Blood samples were analyzed for gamma interferon using a QuantiFERON-TB Gold Plus assay. A complete baseline health profile, including complete blood count, blood glucose, liver and renal functions, and ferritin levels was estimated to identify potential biomarkers that are independently associated with LTBI.

**Results:** The present study revealed an LTBI prevalence of 25.3% (n=99) among PHC workers. The red cell distribution width was significantly associated with LTBI positivity among different biochemical and haematological parameters analyzed. Factors such as individuals’ age, underlying comorbid conditions (30.3%; n=30), and longer employment duration (28%; n=28) were significantly associated with IGRA positivity. IGRA positivity was significantly associated with decreased RDW-SD in females, overweight, and participants with ‘O’ blood group.

**Conclusions:** The study reported a high prevalence of LTBI among HCWs of PHCs, which necessitates their periodical screening for the elimination of TB.

## Introduction

Pulmonary tuberculosis (TB) predominates among the infectious causes of death especially in low- and middle-income countries despite the availability of effective diagnostic tools and anti–tubercular therapeutic (ATT) drugs. The WHO framework with the *End-TB Flagship Strategy* was initiated to reduce TB incidence and deaths.^1^ However, the COVID-19 pandemic appears to have disrupted the critical components of the End–TB strategy – *Find and Treat All*. A significant reduction (∼18%) of newly diagnosed TB was reported in 2020, while a surge in cases and deaths was noticed during 2021–22.^2^ The respiratory system shares the battlefield for both SARS-CoV-2 and *M. tuberculosis* (MTB), which can trigger lung damage.^3^ The increase in TB incidence and deaths post-COVID-19 could be attributed to a perturbed immune system and subsequent reactivation of latent TB infection (LTBI).^4^ Although one–third of the global population remains infected, only ∼5–10% of latently infected reactivate within two years of primary infection.^5^ The latently infected individuals act as a major reservoir from whom reactive TB disease is developed and transmitted in the community, which poses a challenge to TB prevention and treatment. Certain underlying comorbidities including renal and liver diseases, malnutrition, and prolonged exposure to TB-infected individuals likely increase the risk of transition of LTBI into clinical TB disease.^6–8^

The incidence rate of LTBI varies among HCWs ranging from 13–22% who own the highest risk of progression to active TB disease and transmission.^9^ Nonetheless, the prevalence of LTBI among HCWs of PHCs remains largely unknown. LTBI detection by tuberculin skin test (TST) suffers several pitfalls including higher turn-around-time, false-positivity due to the cross-reaction with the vaccine, and other bacterial antigens. It also suffers from poor sensitivity in immunocompromised subjects and in differentiating active disease from latent infections.^10–12^ Implementation of guidelines for TB prevention among HCWs has been one of the top priorities of the National TB Elimination Programme. Therefore, it is imperative to identify and treat appropriately to achieve the goal of ending TB by 2030. Here, a cross-sectional study was carried out on the prevalence of LTBI among primary HCWs in Chennai, India. To assess a more reliable estimate of their relative risk for LTBI, a comprehensive analysis of socio-demographic details, associated risk factors, and clinical and laboratory investigations were carried out.

## Material and Methods

### Study design

The HCWs including physicians, nurses, pharmacists, laboratory technicians, village health nurses, and hospital staff (health assistants, housekeeping, educators, and clerks) from 64 PHCs who maintain close contact with patients with confirmed or unknown TB disease status, were recruited between March and April 2024. Individuals aged ≥18 years, who consented to participate with no clinical evidence of TB were considered. Individuals <18 years of age, pregnancy, HIV infection, and a history of active or latent TB infection were excluded.

A total of 392 individuals meeting the inclusion criteria were recruited into the study. A detailed clinical proforma on clinical history, comorbidities, and risk factors together with sociodemographic details were collected from each participant (Figure 1). The study was approved by the Institutional Ethical Committee.

**Figure 1.**
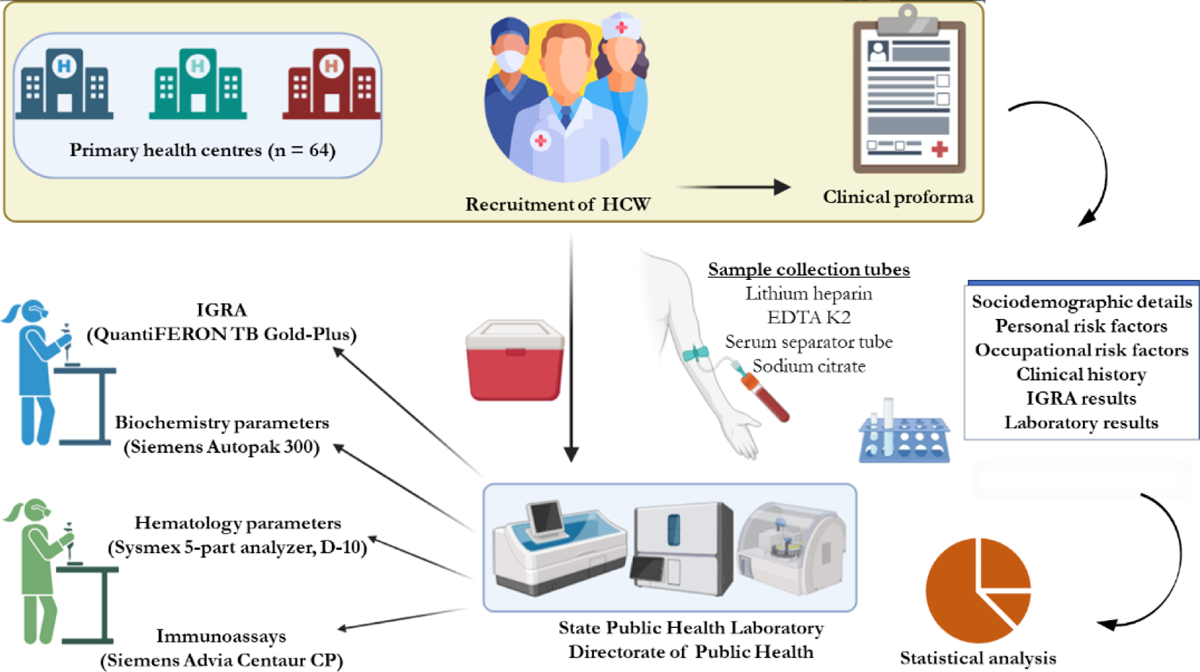
Schematic flowchart of the study.

### Clinical specimens

Blood samples in lithium heparin tubes (5 ml), gel tubes (3 ml), EDTA tubes (2 ml), and citrate tubes (1.6 mL) were collected by venipuncture. One PHC for every administrative block was selected as a collection hub from where the samples were transported in the cold chain to the State Public Health Laboratory (SPHL), Directorate of Public Health and Preventive Medicine within two hours of collection. All the samples were promptly processed for haematological, biochemical, and immunological analyses. Appropriate quality controls were included for all the diagnostic assays.

### Haematological and biochemical parameters

The erythrocyte sedimentation rate (ESR) by Westergren method and complete blood count (CBC) were analyzed using a 5-part haematology analyzer (XN-550, Sysmex). The test parameters included total WBC count, RBC count, haemoglobin, haematocrit (HCT), mean corpuscular volume (MCV), mean corpuscular haemoglobin (MCH) and its concentration (MCHC), platelet count (PLT), red cell distribution width (RDW-SD and RDW-CV), platelet distribution width (PDW), mean platelet volume (MPV), platelet large cell ratio (P-LCR), and plateletcrit (PCT). In addition, differential counts and percentages of neutrophils, lymphocytes, monocytes, eosinophils, and basophils were also assessed. The mean cell size and the breadth of the distribution curve were used to calculate the RDW-CV. The standard deviation (SD) of the mean cell size was divided by MCV to derive the percentage values. The RDW-CV typically ranges between 11 and 15%. Biochemical tests including liver function test, renal function test, glycosylated haemoglobin (HbA1C) as a measure of glucose levels, and C-reactive protein (CRP) were carried out using an automated Autopak 300 clinical chemistry analyzer (Siemens) and a D-10 haemoglobin testing system (Bio-Rad).

### Interferon-gamma release assay (IGRA)

The blood samples were processed for IGRA testing using a commercial QuantiFERON TB-Gold plus assay (Qiagen) as per the manufacturer’s instructions. The test employed a peptide cocktail simulating mycobacterial proteins to stimulate lymphocytes in the heparinized whole blood. The values are expressed in IU/mL. A TB1/TB2 value (minus Nil) of ≥0.35 and ≥25% of Nil was considered positive and reported as a potential MTB infection.

### Ferritin

Ferritin level was determined using the Advia Centaur CP Immunoassay system (Siemens) as per the manufacturer’s instructions. The serum ferritin concentration was expressed as ng/µL. The kit utilized a two-site sandwich chemiluminescence method and had an assay range of 0.5–1650 ng/µL. The normal reference range for males was 22–332 ng/µL and for females was 10–291 ng/µL as recommended by the assay kit.

### Statistical analysis

The primary aim was to report the prevalence of LTBI among HCWs. The association between the LTBI positivity and patient demographics was analyzed using the Chi-square test and Mann-Whitney U test depending on the categorical variable or continuous variable, respectively. Biomarkers were analyzed together using a standard Z-score. Each biomarker was first transformed to Z-score using the following formula, z=(x–μ)/σ, where (x) = subject’s biomarker levels, (μ) = mean of the biomarker levels, and (σ) = standard deviation biomarker levels. The predictive power of biomarkers in aiding the prediction of LTBI-positivity was examined using receiver operating characteristic (ROC) analysis. Fold change was done by normalising the levels of a biomarker in the IGRA-positive group against the median level of the same biomarker in the LTBI-negative group, and the biomarker with significant fold changes was plotted in a heatmap according to the colour scheme indicated. Logistic regression analysis was performed univariately to identify biomarkers that were significantly associated with the LTBI-positivity and plotted as volcanic plots. The biomarkers that showed significant association in the univariate were regarded as candidate predictors and were included in multivariate regression analysis. The odds ratio (OR) and 95% confidence interval (CI) were estimated. Statistical analyses were performed using PRISM software, ver.5.02 (GraphPad Software, San Diego, CA). Logistic regression was performed using SPSS software, ver. 20 (IBM, USA). A p-value of ≤0.05 was considered to be significant. Due to the exploratory nature of the analyses and the dispensable rate of false positives, correction for multiple comparisons was not required.

## Results

A total of 392 individuals between May and June 2024 were included. A detailed clinical proforma was collected from each participant to assess the associated risk factors, and comprehensive laboratory investigations were carried out from the blood samples. The participants from each PHC included different categories of HCWs. Among these, hospital workers were higher in number (n=174; 44.4%) followed by nurses (n=149; 38.0%), and physicians (n=69; 17.6%). The age of the participants ranged from 23 to 60 years with a median of 38 years; The number of female participants was 298 (76%) and male participants was 94 (24.0%). Among nurses, 109 were female (93.0%), among physicians 40 were female (58.0%), among pharmacists, 16 were female (53.0%), and among laboratory technicians, 52 were female (89.6%) (Figure 2).

**Figure 2.**
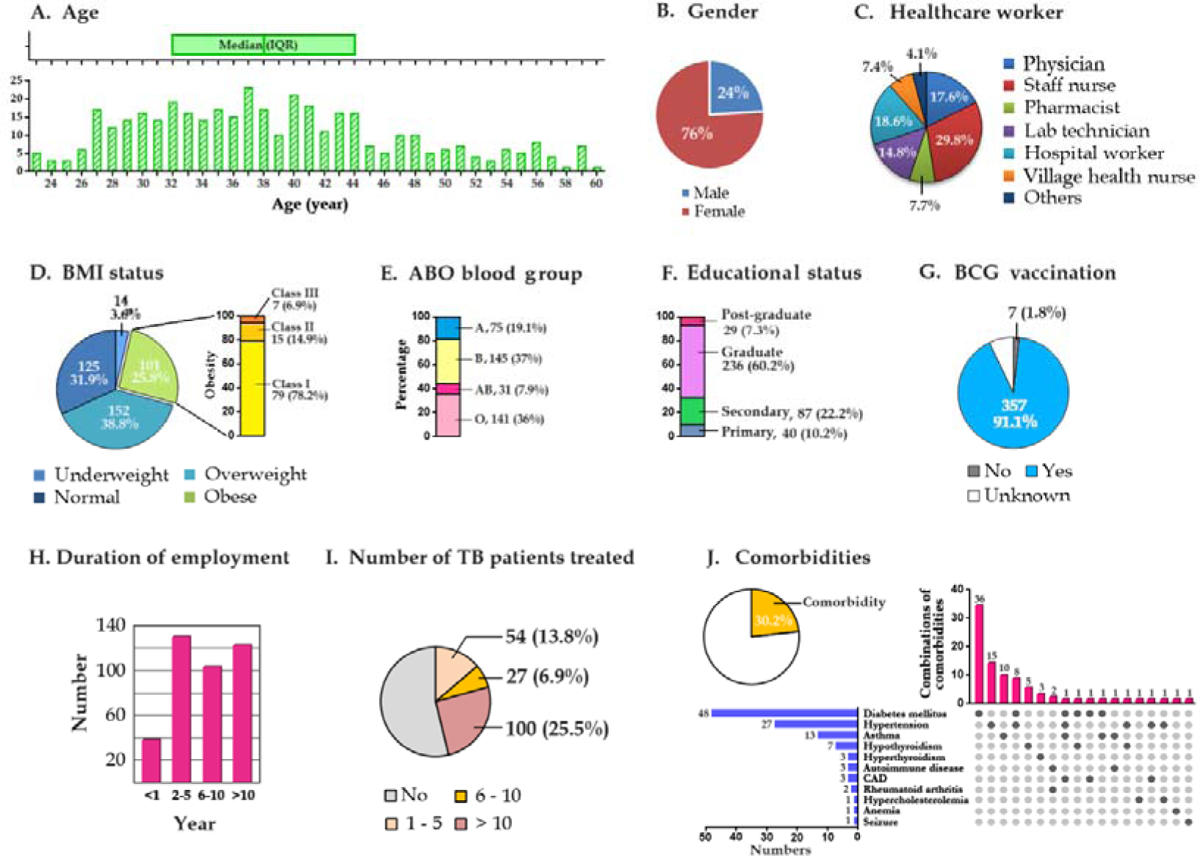
Clinical and demographic description of the study cohort. (A) age distribution of the cohort, (B) gender, (C) healthcare worker, (D) BMI status, (E) ABO blood group, (F) educational status, (G) BCG vaccination status, (H) duration of employment in hospital, (I) number of TB patients treated, (J) comorbidities.

Among the 392 samples tested, 99 (25.3%) revealed a positive IGRA test. Among the different categories of HCWs tested, village health nurses showed the highest positive rate (38.0%) followed by pharmacists (33.0%), hospital workers (32.0%), and staff nurses (25.0%). The percentage of IGRA positives among males (28.7%) did not differ significantly from females (24.2%, p=0.414). The domiciliary status of the study participants among the IGRA-positives was also analyzed. Although participants from rural were higher in numbers (61.5%) compared to urban areas, the percentage of IGRA-positive cases in rural and urban populations were 27.8% and 21.2% respectively and the association between the domiciliary status and IGRA-positive rate was not statistically significant (p=0.153).

Measurement of body mass index (BMI) among the 392 participants indicated that a larger proportion were overweight (38.8%), 3.6% were underweight, and 31.9% showed normal BMI. Intriguingly, 25.8% of individuals were identified with class III obesity. IGRA was highest in class III obese participants (31.0%) followed by participants with normal BMI (25%). While a majority of the participants were vaccinated for Bacillus Calmette-Guérin (BCG) and were aware of their vaccination status (91.1%), 7.1% were unfamiliar with their vaccination history, and 1.8% had not been vaccinated. The number of IGRA positives was higher in the population with unknown vaccination status (32.1%) followed by the vaccinated population (24.9%). Among the HCWs, 60.2% were graduates and 22.2% had completed secondary education. However, educational categories did not show any difference between IGRA positives and negatives. Of the 392 participants, n=21 (5.4%) disclosed a fair control of blood glucose with a mean HbA1c value of 7.4 and a mean blood glucose level of 166 mg/dL. Among the 392 participants, 52.6% (n=206) had abnormal LFT and 25.3% (n=99) had abnormal RFT (Table 1). No statistically significant difference was observed between the two groups. SARS-CoV-2 IgG was detected among all participants except two, of which one was positive for IGRA and the other was negative.

**Table 1.** Sociodemographic and clinical characteristics among IGRA positive individuals.

| **Total sample tested = 392** | **N (%)** | **IGRA Positive N (%)** | **IGRA Negative N (%)** |
| --- | --- | --- | --- |
| Males | 94 (24) | 27 (27.3) | 67 (22.9) |
| Females | 298 (76) | 72 (72.7) | 226 (77.1) |
| **Comorbidities** |  |  |  |
| Asthma | 13 (3.3) | 3 (3) | 10 (3.4) |
| Thyroid disorders | 10 (2.5) | 5 (5) | 5 (5) |
| Diabetes | 48 (12.2) | 15 (15.2) | 33 (11.3) |
| Hypertension | 27 (6.9) | 8 (8.1) | 19 (6.5) |
| Anemia | 1 (0.3) | 1 (1) | 0 |
| Autoimmune disease | 3 (0.8) | 1 (1) | 2 (0.7) |
| Seizures | 1 (0.3) | 1 (1) | 0 |
| None | 289 (73.7) | 65 (65.7) | 224 (76.5) |
| **Domiciliary status** |  |  |  |
| Rural | 241 (61.5) | 67 (67.7) | 174 (59.4) |
| Urban | 151 (38.5) | 32 (32.3) | 119 (40.6) |
| **BMI** |  |  |  |
| Underweight | 14 (3.6) | 2 (2) | 12 (4.1) |
| Normal | 125 (31.9) | 32 (32.3) | 93 (31.7) |
| Obese class 3 | 101 (25.8) | 32 (32.3) | 69 (23.5) |
| Overweight | 152 (38.8) | 33 (33.3) | 119 (40.6) |
| **BCG** |  |  |  |
| Vaccinated | 357 (91.1) | 89 (89.9) | 268 (91.5) |
| Unknown | 28 (7.1) | 9 (9.1) | 19 (6.5) |
| Not vaccinated | 7 (1.8) | 1 (1) | 6 (2) |
| **Education** |  |  |  |
| Primary | 40 (10.2) | 13 (13.1) | 27 (9.2) |
| Secondary | 87 (22.2) | 24 (24.2) | 63 (21.5) |
| Graduate | 236 (60.2) | 53 (53.5) | 183 (62.4) |
| Post-graduate | 29 (7.4) | 9 (9.1) | 20 (6.8) |
| **Work experience** |  |  |  |
| Less than a year | 39 (9.9) | 6 (6.1) | 33 (11.3) |
| 2 to 5 | 128 (32.7) | 25 (25.3) | 103 (35.1) |
| 6 to 10 | 103 (26.3) | 34 (34.3) | 69 (23.5) |
| More than 10 years | 122 (31.1) | 34 (34.3) | 88 (30) |
| **TST** |  |  |  |
| Done | 20 (5.1) | 6 (6.1) | 14 (4.8) |
| Not done | 372 (94.9) | 93 (93.9) | 279 (95.2) |
| **Treated TB case** |  |  |  |
| 1 to 5 cases | 54 (13.8) | 9 (9.1) | 45 (15.3) |
| 6 to 10 cases | 27 (6.9) | 7 (7.1) | 20 (6.8) |
| More than 10 cases | 100 (25.5) | 27 (27.2) | 73 (24.9) |
| No | 211 (53.8) | 56 (56.5) | 155 (52.9) |
| **HbA1C** |  |  |  |
| Fair control | 21 (5.4) | 9 (9.1) | 12 (4.1) |
| Good control | 82 (20.9) | 19 (19.2) | 63 (21.5) |
| Normal | 250 (63.8) | 58 (58.6) | 192 (65.5) |
| Poor control | 39 (9.9) | 13 (13.1) | 26 (8.9) |
| **LFT** |  |  |  |
| Abnormal | 206 (52.6) | 51 (51.5) | 155 (52.9) |
| Normal | 186 (47.4) | 48 (48.5) | 138 (47.1) |
| **RFT** |  |  |  |
| Abnormal | 99 (25.3) | 19 (19.2) | 80 (27.3) |
| Normal | 293 (74.7) | 80 (80.8) | 213 (72.7) |
| **CRP** |  |  |  |
| Abnormal | 114 (29.1) | 35 (35.4) | 79 (26.9) |
| Normal | 278 (70.9) | 64 (64.6) | 214 (73) |
| **Smoking** |  |  |  |
| Yes | 4 (1) | 2 (2) | 2 (0.7) |
| No | 388 (99) | 97 (97.9) | 291 (99.3) |
| **Drinking** |  |  |  |
| Yes | 14 (3.6) | 4 (4) | 10 (3.4) |
| No | 378 (96.4) | 95 (95.9) | 283 (96.6) |
| **Anti-SARS-CoV-2 IgG** |  |  |  |
| Reactive | 390 (99.5) | 98 (98.9) | 292 (99.6) |
| Non-reactive | 2 (0.5) | 1 (1) | 1 (0.3) |
| **Ferritin** |  |  |  |
| Abnormal | 57 (14.5) | 13 (13.1) | 44 (15) |
| Normal | 335 (85.5) | 86 (86.9) | 249 (84.9) |

Among different demographic markers analyzed, the age of the participants and duration of employment were significantly associated with IGRA positivity (p=0.029 and p=0.034, respectively); however, gender and BMI were not significantly associated. Interestingly, neither TB exposure history nor the number of TB cases treated by the HCW had a significant association with LTBI positivity indicating a possible acquisition of infection from the hospital but likely not directly from treated patients.

To improve comparability between biomarkers with different scales, the level of each biomarker was transformed into a uniform scale, the Z-score. When compared between IGRA-positives and -negatives, the positives showed a statistically significant lower Z-score in RDW-SD, RDW-CV% (p=0.002 and p=0.026, respectively), and had a higher Z-score in eosinophil percentage (p=0.031) (Figure 3A). ROC analysis was performed between IGRA-positive and -negative participants to assess the suitability of RDW-SD, RDW-CV%, and eosinophil percentage as surrogate markers of LTBI. The diagnosis efficacy of RDW-SD and eosinophil percentage for diagnosis of LTBI was only marginal, where the AUC of 0.574; p=0.002 and AUC=0.597; p=0.004, respectively. The AUC for RDW-CV% was not significant (Figure 3B).

**Figure 3.**
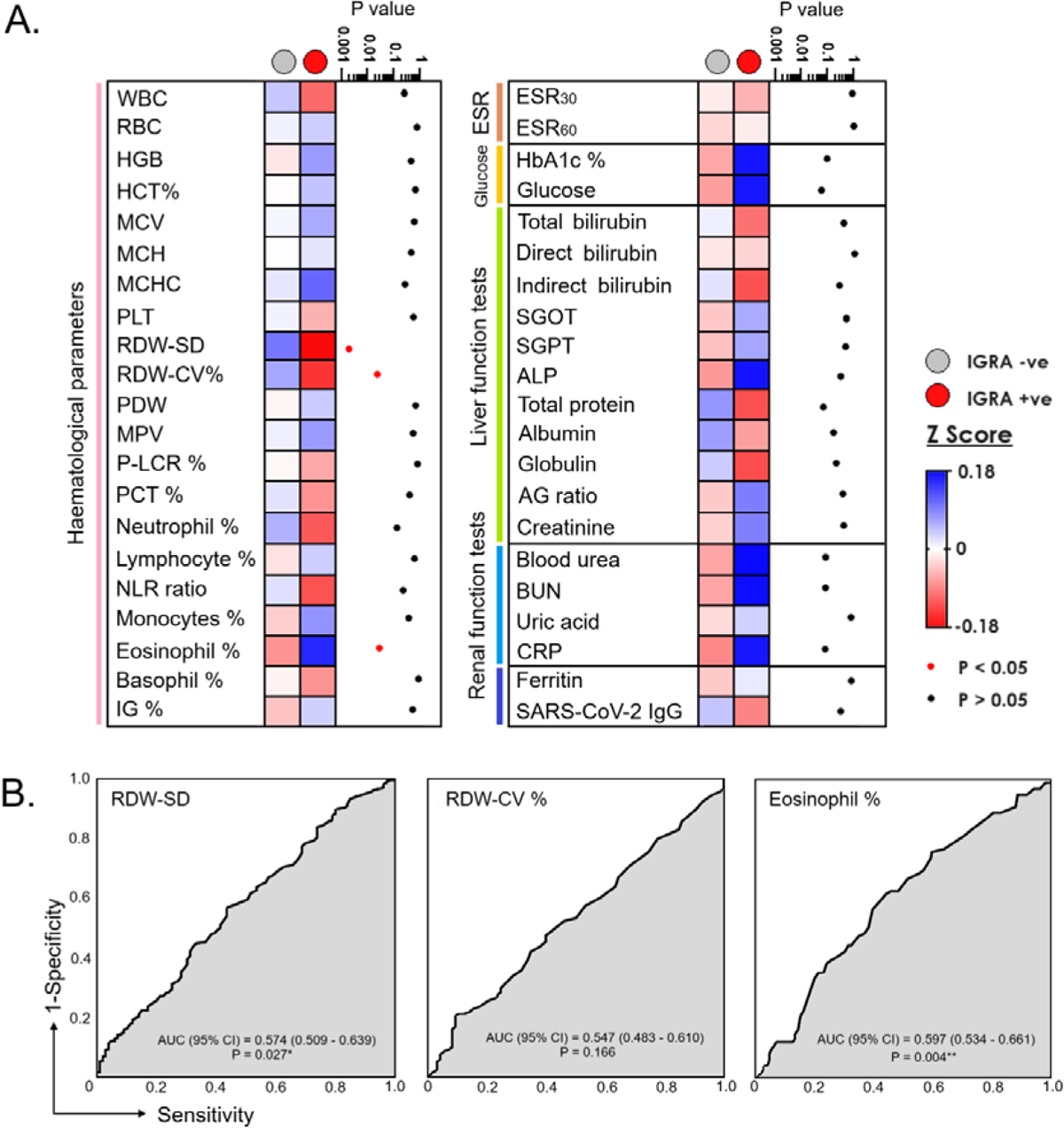
(A) Comparison of the standardized levels (Z-score) of parameters among HCWs with IGRA+ve and IGRA-ve (B) Receiver operating characteristic (ROC) curves for prediction of latent tuberculosis infection (LTBI) using RDW-SD, RDW-CV%, and eosinophils %; AUC = Area under curve. * and ** represent *p<*0.05, and *<*0.01, respectively.

Consummated from our previous studies on biomarkers such as plasma CXCL8 and MCP-1 that aid in diagnosis and as surrogate predictors of disease progression^13^ and platelet-large cell ratio and ESR^14^ that were probably affected by demographics, we here measured the fold change of each biomarker (normalized against IGRA-negative) and stratified under gender, ABO blood grouping, and BMI status. Biomarkers that showed significant alteration in fold change were indicated according to the colour scheme (Figure 4A). The results showed that the levels of biomarkers were differentially altered in gender, BMI status, and ABO blood grouping. The results showed that IGRA positivity was significantly associated with decreased RDW-SD in females, overweight, and participants with the ‘O’ blood group. Eosinophils percentage was significantly associated with IGRA positivity in females, participants with normal weight and obesity as well as participants with blood group type ‘B’. Participants with blood group ‘O’ showed the most profound alteration in haematological markers, where IGRA positivity was associated with decreased percentages of neutrophils and immunoglobulin, as well as increased percentages of lymphocytes and monocytes. Besides, participants with IGRA positivity also showed elevation of blood urea and BUN in total participants, females, participants with normal weight as well as participants with blood groups B and AB.

**Figure 4.**
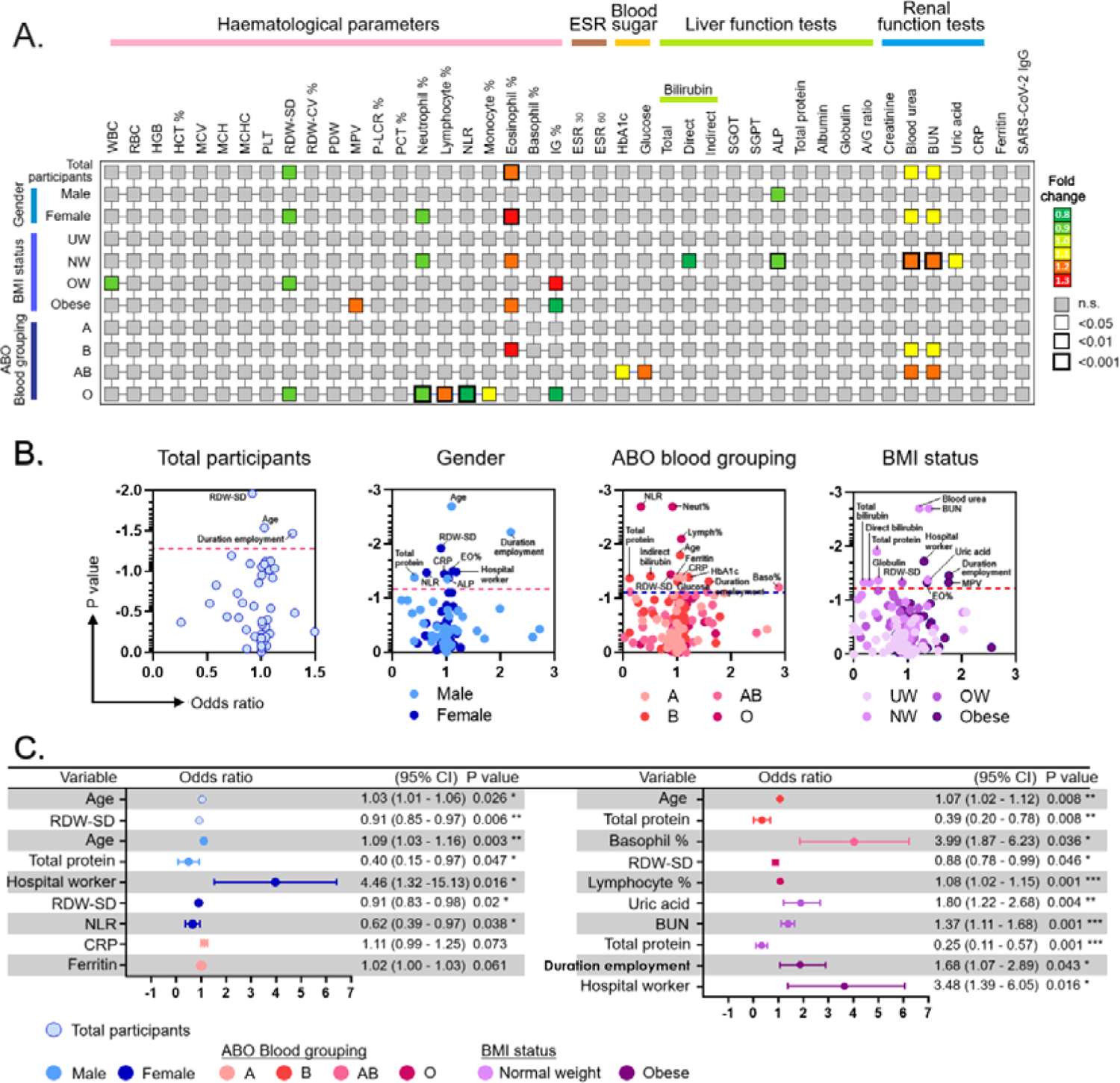
Parameters associated with LTBI among HCWs stratified by gender, BMI status, and ABO blood group. (A) Comparison of fold-change of parameters and their association with LTBI, (B) Univariate logistic regression analysis of parameters associated with LTBI, (C) Multivariate logistic regression analysis of parameters, associated with LTBI (The Hosmer–Lemeshow value for this model was P=0.162).

To identify the biomarker that is independently associated with IGRA-positivity, we performed logistic regression analysis whereby each biomarker was tested univariately against IGRA-positive, and biomarkers with p<0.05 were regarded as candidates (Figure 4B). The candidates were then subjected to a multivariate logistic regression analysis and biomarkers with p*<*0.05 were considered as independent predictors in their respective models (Figure 4C). We found that age was significantly associated with increased odds of IGRA-positive, whereby every increase of age by one year was associated with increased odds of having IGRA-positive by 3.0%, (95% CI = 1–6%); p=0.026. Whilst every decrease of RDW-SD by one unit was significantly associated with decreased odds of IGRA-positive by 9%, (95% CI = 3–15%); p=0.006.

Considering age, BMI, and ferritin as the confounding factors for CRP, logistic regression analysis indicated a significant association of age-adjusted CRP (p=0.026) and age-adjusted ferritin (p=0.032) with LTBI-positive individuals. The univariate and multivariate logistic regression analyses on each parameter were carried out (Figure 4c). Spearman’s correlation (rho) was used to evaluate the relationship between the biomarkers and determine the biomarkers that are in colinearity. The correlations were considered significantly strong if rho≥ 0.60 and p <0.05. The study did not show any biomarkers with strong colinearity in the logistic regression model (see Supplementary Figure 1).

## Discussion

We carried out a cross-sectional study to estimate the prevalence of LTBI among HCWs of primary health centres across Chennai, India. With a prevalence of 25.3% among the healthcare workers, the highest positivity rate was observed with village health nurses followed by other workers including staff nurses and physicians.

For effective control of TB, appropriate identification and treatment of latently infected individuals are vital to reduce their risk of progression to active disease, especially among the risk groups. PHCs in India act as the frontline in medical care for the local population, particularly at the community level, and provide affordable healthcare services including laboratory diagnosis and treatment towards universal health coverage.^15^ HCWs are considered to be the moderate-risk group for TB reactivation. Other factors such as malnutrition, immune suppression or weakened immune system, smoking, and comorbidities such as diabetes, thyroid dysfunction, renal dysfunction, and liver dysfunction may increase the risk substantially.^16–21^ In addition, increased tuberculosis is contributed by social or environmental determinants and personal health determinants which are often ignored while assessing the prevalence of LTBI in the community. Overall, this puts the determination of universal health coverage in jeopardy when the fundamental healthcare system at the primary level is affected. In our study, we determined to study the prevalence of LTBI among HCWs of PHCs and assess their LTBI-associated laboratory parameters using multiple logistic regression analysis.

Our previous investigation on household contacts indicated a 43% positive rate with a female predominance.^13^ It is therefore quintessential to identify LTBI test-positive individuals who act as reservoirs and seedbeds for transmission of reactive TB disease in the community. Likewise, HCWs have a very high occupational risk of latent infection and are susceptible to progression to active TB.^22^ However, there are very limited reports on the HCWs especially from PHC. Hence, it is of paramount importance for frontline healthcare providers in low or limited-resource settings to undergo periodical clinical and radiological screening and monitoring of fatal infections like tuberculosis. This is the first study to our knowledge that reports the prevalence of LTBI and associated risk factors among HCWs of primary health centres. The present study reported 25.3% positivity similar to trends seen among other risk groups such as household contacts.^23^

LTBI prevalence has been shown to increase with increasing duration of exposure.^24^ Conceivably, the inclusion of factors such as exposure proximity, duration, and number of exposures, number of patient contacts could have provided better insights and clinical acumen but was unavailable and therefore not analyzed in this study. Mostly, individuals who show a positive IGRA result may show a positive result throughout their lifetime despite the use of anti-tubercular drugs. The IGRA test cannot identify re-infection post-treatment, besides, all LTBI-positive individuals should be screened for active disease as the LTBI treatment regimen could lead to drug resistance. It is therefore important to evaluate the IGRA results carefully considering the patient’s past clinical history. Of our test population (n=392), none of them had a history of TB disease and a total of 20 (5.1%) had a history of TST done previously. Based on their self-declaration, all were negative on testing although the exact timelines of their previous test are unknown. This also implies that these individuals most likely acquired the infection in the healthcare facility recently. It is imperative to have a deeper understanding of the mode of pathogen transmission for better clinical management and to develop appropriate public health policy.

Before the decision whether to treat or not treat LTBI is made, it is important to assess the underlying conditions such as HIV, liver disease, alcohol use, pregnancy, and use of specific medications that could compromise liver function. In our study, all the individuals were not tested for HIV and hepatitis, however, baseline laboratory liver function test and renal function test along with complete blood count were carried out to assess their health. Among 392 tested, 52% had abnormal liver function of which 25% of individuals were IGRA positive. Also, 25% had abnormal renal function, of which 19% were IGRA positive. Although the mean values of renal and liver biomarkers did not differ, the LTBI-positive individuals with abnormal LFT and/or RFT could have an increased risk of disease progression. Nevertheless, the baseline abnormal liver function tests are considered to be a major risk for drug-induced hepatotoxicity when treated for LTBI.^25^ In our study group, 14 (3.6%) had a history of alcohol use, of these 11 continued to alcohol use. Among the 14 alcohol users, four were positive for LTBI and all were current alcohol users. Alcohol use is a major risk factor for a weakened immune system and substantially increases the risk of reactivation.^26,27^ While an intake of alcohol in moderate quantities is protective against active TB among non-smokers, there is a linear increased risk of disease progression with increased consumption.^28,29^

Four individuals were using immunosuppressive drugs, had no other risk factors and all of them were negative for LTBI. Various immunosuppressants including methotrexate, prednisolone, and phenytoin were found to be being used in these individuals. The use of immunosuppressive drugs other than reactivation of latent TB has also been shown to impact the latent TB testing, however, IGRA tests are shown to be more sensitive than TST for immunosuppressed individuals. IGRA assay in contrast to skin testing, does not have a predetermined cut-off value and is specific to mycobacterial antigens. The proposed guideline indicates preventive treatment for LTBI without further testing for high-risk groups.^30^ The prevalence of LTBI among pregnant women ranges from 14 to 48% in the USA and shows an increased risk of reactivation during the postpartum period.^31^ There were no pregnant women in our study population; however, there were five lactating mothers and all five were negative for LTBI. IGRA assay is also reported to be more specific, sensitive, and unaltered by pregnancy.^31^

Among the different haematological parameters tested including absolute count, RDW-CD, RDW-SD, PDW, PCT, MPV, and platelet-large cell ratio, RDW-CV was found to be highly significant in LTBI-positive individuals. The significance of haematological markers such as RDW-CV, RDW-SD, and inflammatory markers such as CRP have been shown to correlate with the severity and prognosis of different diseases including TB.^32–35^ This is the first report on the use of RDW-CV and RDW-SD as significant markers indicative of LTBI. Considering the cut-off limits of RDW-CV, among 99 LTBI positives, 15.0% had abnormal RDW and among 293 LTBI negatives, 24.0% had abnormal RDW. Its association and usefulness in differentiating LTBI should be further investigated with a large sample size and heterogenous test population. Likewise, ferritin level is an important factor when stratified with age, sex, CRP, and other inflammatory markers in determining the magnitude and distribution of iron deficiency and iron overload as a public health problem. Different studies indicated their role in predicting LTBI and prognosis.^36,37^ Serum ferritin levels along with other inflammatory biomarkers such as CRP were found in two studies to indicate mycobacterial load and TB-associated inflammation during anti-tuberculous therapy.^38,39^ Another study showed many markers including serum ferritin and certain risk factors not significantly associated with LTBI individuals.^40^ Yet, iron metabolism indices could be further evaluated with different populations to evaluate its efficacy in differentiating LTBI from active disease and in prognosis.

LTBI among HCWs, especially in resource-limited settings is of major global concern. The healthcare workforce striving towards ending TB is largely affected due to their occupational risk. Hence, routine screening and appropriate medical interventions are crucial for the successful management of TB disease. Coordinated scientific efforts and clinical strategies with appropriate aligned public health measures are urgently warranted for TB elimination.

## Supplementary Files

Supplementary Figure 1. Spearman correlation analysis between the estimated biomarkers.

## Ethical approval statement

The study was approved by the Institutional Ethical Committee (Approval Reference number: DPHPM/IEC/010/V2 Dated 20/03/2024). Participants provided written informed consent themselves for the study.

## Author contributions

Sivaprakasam T. Selvavinayagam, Masilamani Senthilkumar, Marie Larsson, Pachamuthu Balakrishnan, Vijayakumar Velu, Esaki M. Shankar, and Sivadoss Raju designed the study and were responsible for conceptualization and data curation. Sivaprakasam T. Selvavinayagam, Sivadoss Raju, Esaki M. Shankar, Marie Larsson, Vijayakumar Velu, Pachamuthu Balakrishnan, Ganga Sankar, Sathish Sankar, Yean K. Yong, Hong Y. Tan, Ying Zhang, Manivannan Rajeshkumar, Nagarajan Muruganandam, Deepak M. Sakate, Deepak Jayakumar and Prabu Dhandapani conducted the analyses and were responsible for methodology, formal analysis, validation, and visualization. Asha Frederick, Paulkanraj PriyaRaj, Jesudoss Prabhakaran, Pattusamy Sangeetha, Pasupathy Arunpathy, Rajamani Charu were responsible for recruitment of participants, data collection, analysis and data curation. Sivaprakasam T. Selvavinayagam, Sathish Sankar, Marie Larsson, Vijayakumar Velu, Esaki M. Shankar, and Sivadoss Raju wrote the first draft of the manuscript. All authors provided critical inputs and approved the final version of the manuscript for publication.

## Data Availability

All data produced in the present work are contained in the manuscript.

---

**Supplementary Figure 1.**
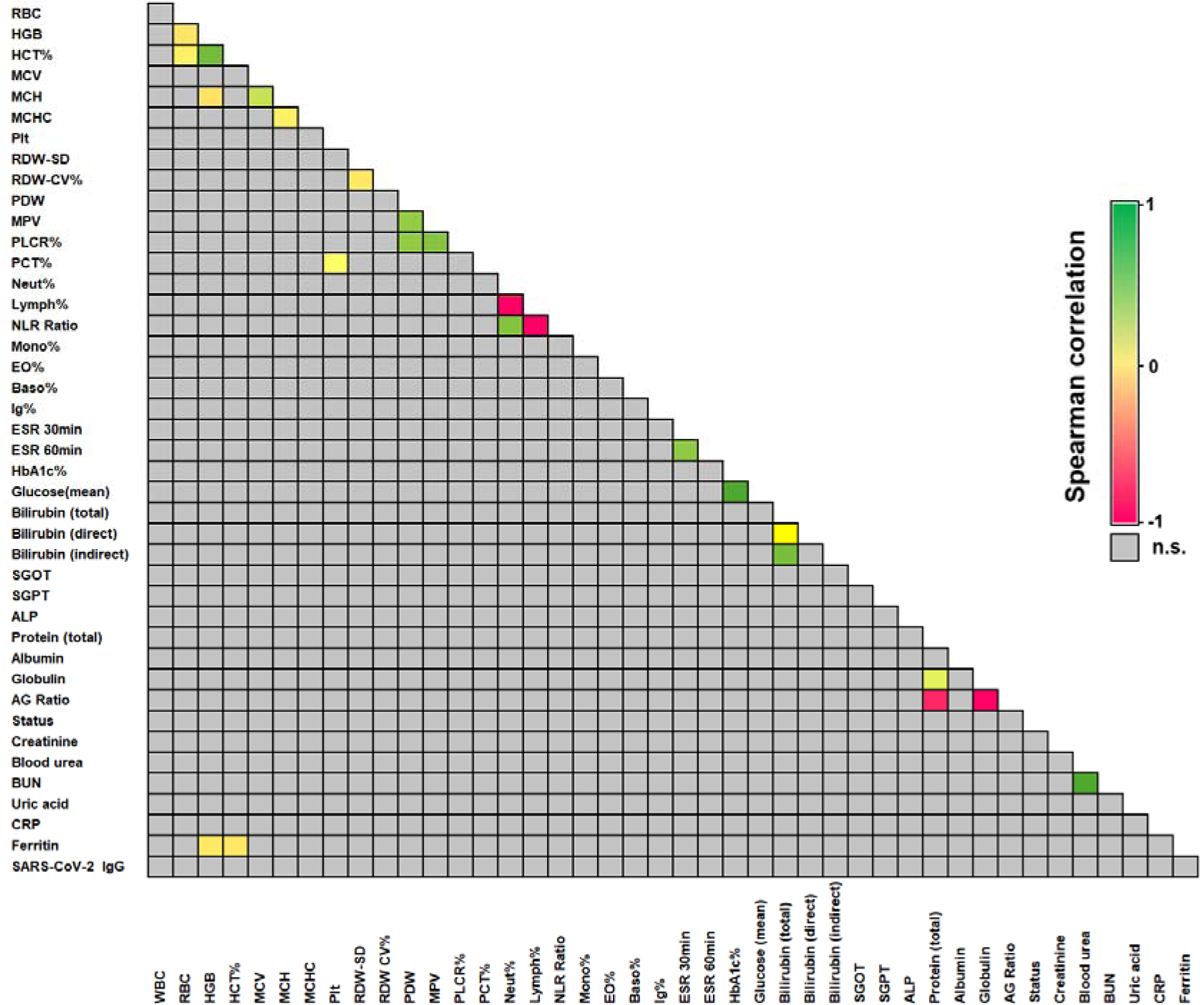
Spearman correlation analysis between the estimated biomarkers.

