## Supplementary figures and images for "Burden of Latent Tuberculosis Infection among Healthcare Workers of Primary Health Centres and its Association with Clinical Laboratory Parameters"

### Supplemental Figure 1

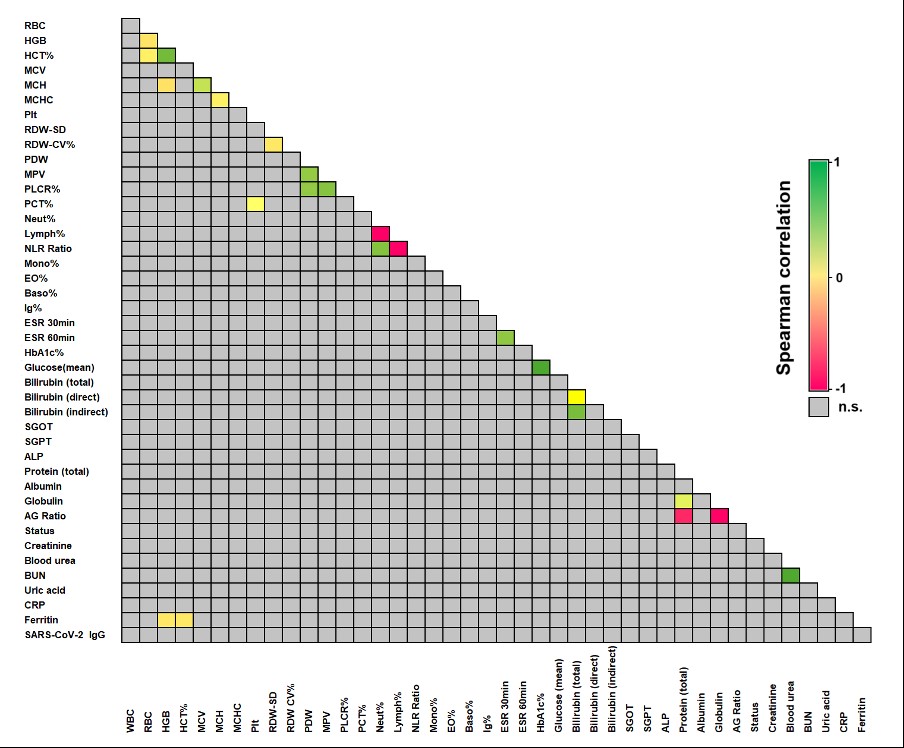
